# Gastroesophageal reflux disease is associated with differences in the allograft microbiome, microbial density and inflammation in lung transplantation

**DOI:** 10.1101/2021.09.03.21263067

**Authors:** Pierre H.H. Schneeberger, Chen Yang Kevin Zhang, Jessica Santilli, Bo Chen, Wei Xu, Youngho Lee, Zonelle Wijesinha, Elaine Reguera-Nuñez, Noelle Yee, Musawir Ahmed, Kristen Boonstra, Rayoun Ramendra, Courtney W. Frankel, Scott M. Palmer, Jamie L. Todd, Tereza Martinu, Bryan Coburn

## Abstract

**Rationale:** Gastroesophageal reflux disease (GERD) may affect lung allograft inflammation and function through its effects on allograft microbial community composition in lung transplant recipients.

**Objectives:** Our objective was to compare the allograft microbiota in lung transplant recipients with or without clinically diagnosed GERD in the first post-transplant year, and assess associations between GERD, allograft microbiota, inflammation and acute and chronic lung allograft dysfunction (ALAD/CLAD).

**Methods:** 268 bronchoalveolar lavage samples were collected from 75 lung transplant recipients at a single transplant centre every 3 months post-transplant for 1 year. Ten transplant recipients from a separate transplant centre provided samples pre/post-anti-reflux Nissen fundoplication surgery. Microbial community composition and density were measured using 16S rRNA gene sequencing and qPCR, respectively and inflammatory markers and bile acids were quantified.

**Measurements and Main Results:** We observed three community composition profiles (labelled community state types, CSTs 1-3). Transplant recipients with GERD were more likely to have CST1, characterized by high bacterial density and relative abundance of the oropharyngeal colonizing genera *Prevotella* and *Veillonella*. GERD was associated with more frequent transition to CST1. CST1 was associated with lower per-bacteria inflammatory cytokine levels than the pathogen-dominated CST3. Time-dependant models revealed associations between CST3 and development of ALAD/CLAD. Nissen fundoplication decreased bacterial load and pro-inflammatory cytokines.

**Conclusion:** GERD was associated with a high bacterial density, *Prevotella/Veillonella* dominated CST1. CST3, but not CST1 or GERD, was associated with inflammation and early development of ALAD/CLAD. Nissen fundoplication was associated with decreases in microbial density in BALF samples, especially the CST1-specific genus, *Prevotella*.

## Introduction

Gastroesophageal reflux disease (GERD) is common after lung transplantation and is characterized by reflux of gastric contents, including gastric acid, mucus, digestive enzymes and bile acids. This refluxate can be aspirated by some patients, leading to lung allograft injury and inflammation. Indeed, we recently reported an association between GERD, levels of taurocholic acid (TCA, a bile acid) and inflammatory markers in the bronchoalveolar lavage fluid (BALF), and acute lung allograft dysfunction (ALAD) at 3-months post-transplant (1). The association of GERD and chronic lung allograft dysfunction (CLAD) – the leading cause of death in the late post-transplant period (2) – is inconsistent, however, with some, but not all studies indicating GERD is a risk factor for CLAD (3, 4).

The composition of microbial communities in the lung allograft after transplantation varies between individuals and is associated both with CLAD (5, 6) and acute inflammation (7, 8). Composition of the allograft microbial community may follow dynamics similar to those presented in the ecological concept of island biogeography (9) in which community composition is the product of immigration and extinction rates (10, 11). These rates can be affected by multiple factors including individual characteristics such as proximity of the trachea to the oropharynx, and, potentially, GERD.

The composition of allograft microbial communities is associated with distinct patterns of soluble biomarkers in BALF, with communities dominated by *Proteobacteria* or *Firmicutes* being associated with inflammation and *Bacteroidetes* domination associated with markers of airway remodelling (7, 8). We reasoned that allograft microbial community composition, GERD and inflammation may be associated in the post-transplant period, and that models incorporating both GERD and community composition may predict inflammation, ALAD and CLAD better than models including only individual predictors.

To address this, we compared lung allograft microbial community composition from BALF between individuals with and without GERD in the first year after transplant and assessed GERD/microbial-community/inflammation associations, including longitudinal comparisons. We assessed GERD and microbial community composition as predictors of inflammation, ALAD, and CLAD in this cohort. Finally, we measured changes in microbial density and inflammatory cytokines levels before and after Nissen fundoplication in a secondary cohort of lung transplant recipients to determine whether surgical treatment of GERD altered microbial density and inflammation.

## Results

### Cohort features

Baseline patient characteristics of the GERD cohort were comparable between GERD and no-GERD patients (**Table 1**) and inclusion/exclusion criteria are reported in **Figure S1**. Transbronchial biopsy pathology results (A-grades and B-grades), BALF culture results, CLAD diagnoses, and follow-up times are reported for each patient in **Figure S2**. Baseline characteristics of Nissen cohort patients are reported in **Table 2**.

**Table 1.** Baseline patient characteristics of the main cohort.

| Characteristic | GERD<br>(n=24) | no-GERD<br>(n=51) | P value |
| --- | --- | --- | --- |
| **Recipient age at transplant** |  |  | 0.23 |
| Mean ± SD | 54 ± 14 | 57 ± 12 | |
| Range | 19–70 | 22–75 |  |
| Male, n (%) | 14 (58) | 29 (57) | 1.00 |
| **Native lung disease, n (%)** |  |  | 0.98 |
| Chronic Obstructive Pulmonary Disease | 4 (17) | 10 (20) |  |
| Cystic Fibrosis | 2 (8) | 6 (12) |  |
| Pulmonary Fibrosis | 12 (50) | 24 (47) |  |
| Other | 6 (25) | 11 (22) |  |
| **Transplant type, n (%)** |  |  | 1.00 |
| Single lung | 3 (13) | 7 (14) |  |
| Double lung | 24 (87) | 44 (86) |  |
| **Donor recipient CMV status, n (%)** |  |  | 0.85 |
| **D-/R-** | 3 (13) | 11 (22) |  |
| **D-/R+** | 7 (29) | 14 (27) |  |
| **D+/R-** | 6 (25) | 12 (24) |  |
| **D+/R+** | 8 (33) | 14 (27) |  |
| **BAL samples available by time point, n** |  |  |  |
| **3 months** | 23 | 51 |  |
| **6 months** | 20 | 43 |  |
| **9 months** | 20 | 49 |  |
| **12 months** | 19 | 45 |  |
| **24h pH impedance testing, n** |  |  |  |
| **Total reflux episodes** | 8 (5-15) | 66 (57-71) | <0.001 |
| **Proximal reflux episodes** | 5 (2-8) | 38 (26-54) | <0.001 |

**Table 2.** Characteristics of patients undergoing Nissen fundoplication to treat GERD.

| Characteristic | Nissen cohort (n=10) |
| --- | --- |
| Recipient age at transplant |  |
| Mean ± SD | 47 ± 14 |
| Range | 18–65 |
| Male, n (%) | 2 (20) |
| Native lung disease, n (%) |  |
| Chronic Obstructive Pulmonary Disease | 3 (30) |
| Cystic Fibrosis | 3 (30) |
| Pulmonary Fibrosis | 0 (0) |
| Other | 4 (40) |
| Days from transplant to Nissen, mean ± SD | 70 ± 29 |

### Comparison of bacterial community composition in BALF between lung transplant recipients with and without GERD

We first assessed bacterial community composition by 16S rRNA gene sequencing (**Figure 1**). Using a Bray-Curtis dissimilarity matrix, we identified three distinct underlying community clusters, or community state types 1-3 (CST1-3, **Figure 1A**). GERD cases and no-GERD controls had distinguishable community composition distributions (PERMANOVA *P* < 0.001, **Figure 1B**) but GERD status explained only a small amount of compositional variance (R^2^ = 0.013). The proportion of GERD cases with CST1 was greater than that of controls, while the proportion of CST2 was less than controls (*P* = 0.014, OR for CST1 [95% CI] = 2.4 [1.2-4.4]) but a similar proportion of patients in both groups presented CST3 (*P* = 0.12, OR for CST1 [95% CI] = 1.7 [0.9-3.2], **Figure 1C**).

**Figure 1.**
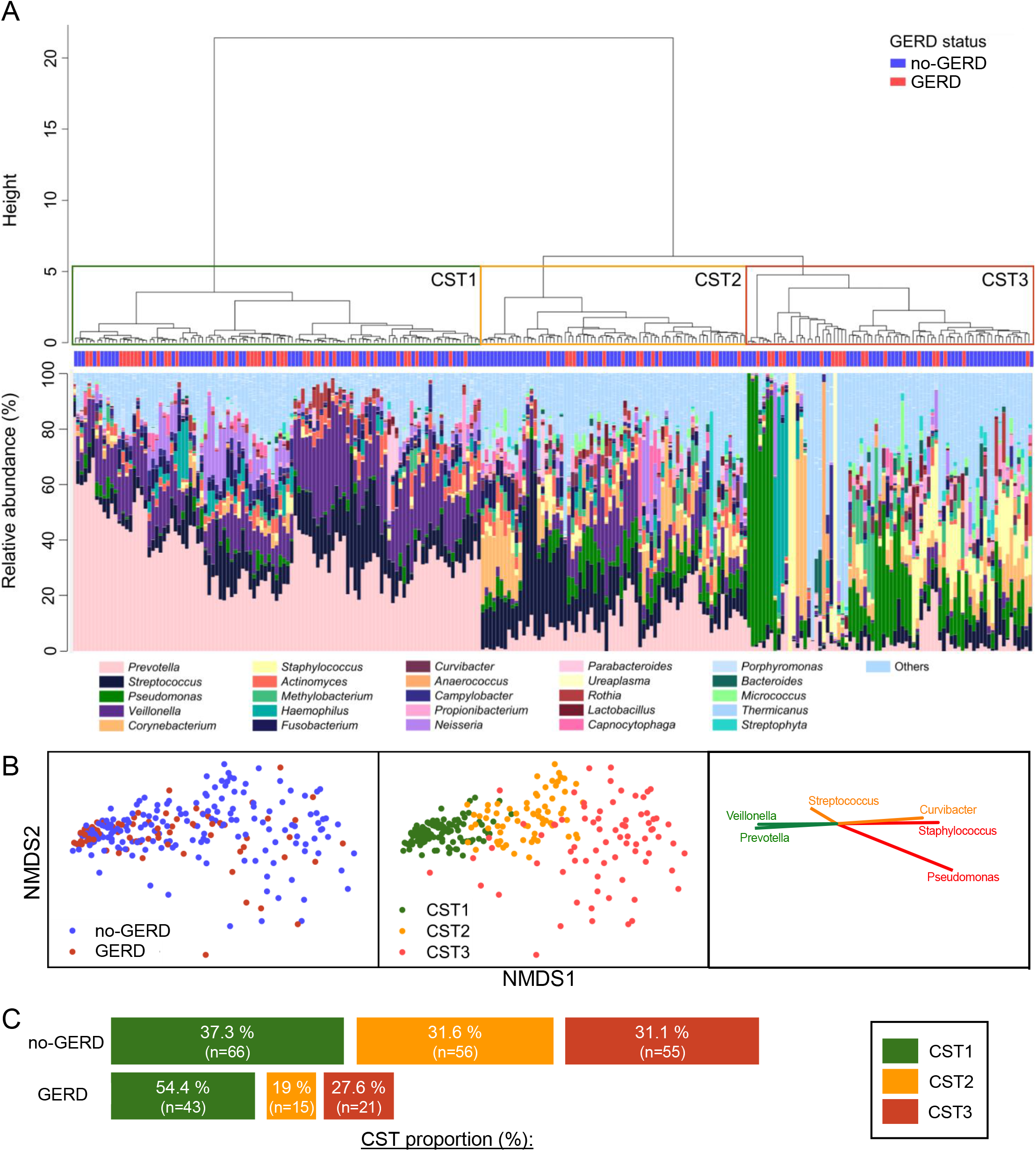
Composition of the lung microbiota in post-transplant patients with and without GERD. **A**. Classification of BALF samples based on Bray-Curtis dissimilarity measure. **B**. Non-metric multidimensional scaling plots based on Bray-Curtis dissimilarity measure comparing spatial ordination of GERD vs no-GERD patients, CST1-3 samples, and showing genera contributing most to this classification system. **C**. Proportion of CSTs by GERD status. GERD = gastroesophageal reflux disease; BALF = bronchoalveolar lavage fluid; CST (1-3) = community state types (1 to 3).

#### Association with Bile Acids

As previously reported (1), TCA was significantly associated with GERD at 3 months post-transplant in this cohort. In this present study, we assessed CST and bile acid associations at 3 months post-transplant. The CSTs were not significantly associated with bile acid concentrations, but weakly trend towards greater TCA and GCA concentrations in CST1 and CST3 (**Figure S3**).

Microbial community composition, bacterial density and within-sample (alpha) diversity differed by CST (**Figure 2**). CST1 was characterized by the high relative abundance of oropharyngeal taxa including *Prevotella* and *Veillonella*. The genera *Streptococcus* and *Tanerella*, were significantly enriched in CST2, while CST3 was characterized by an enrichment of genera with commonly pathogenic species *Pseudomonas* and *Staphylococcus* (**Figure 2A**). The BALF CSTs were distinguished most strongly by the genera *Prevotella, Veillonella, Streptococcus, Pseudomonas*, and *Staphylococcus* with mean decreases of classification accuracy of 0.102, 0.041, 0.025, 0.02, and 0.018, in a random forest model that omitted these taxa, respectively (model classification accuracy=82% and Cohen’s Kappa=73.9%).

**Figure 2.**
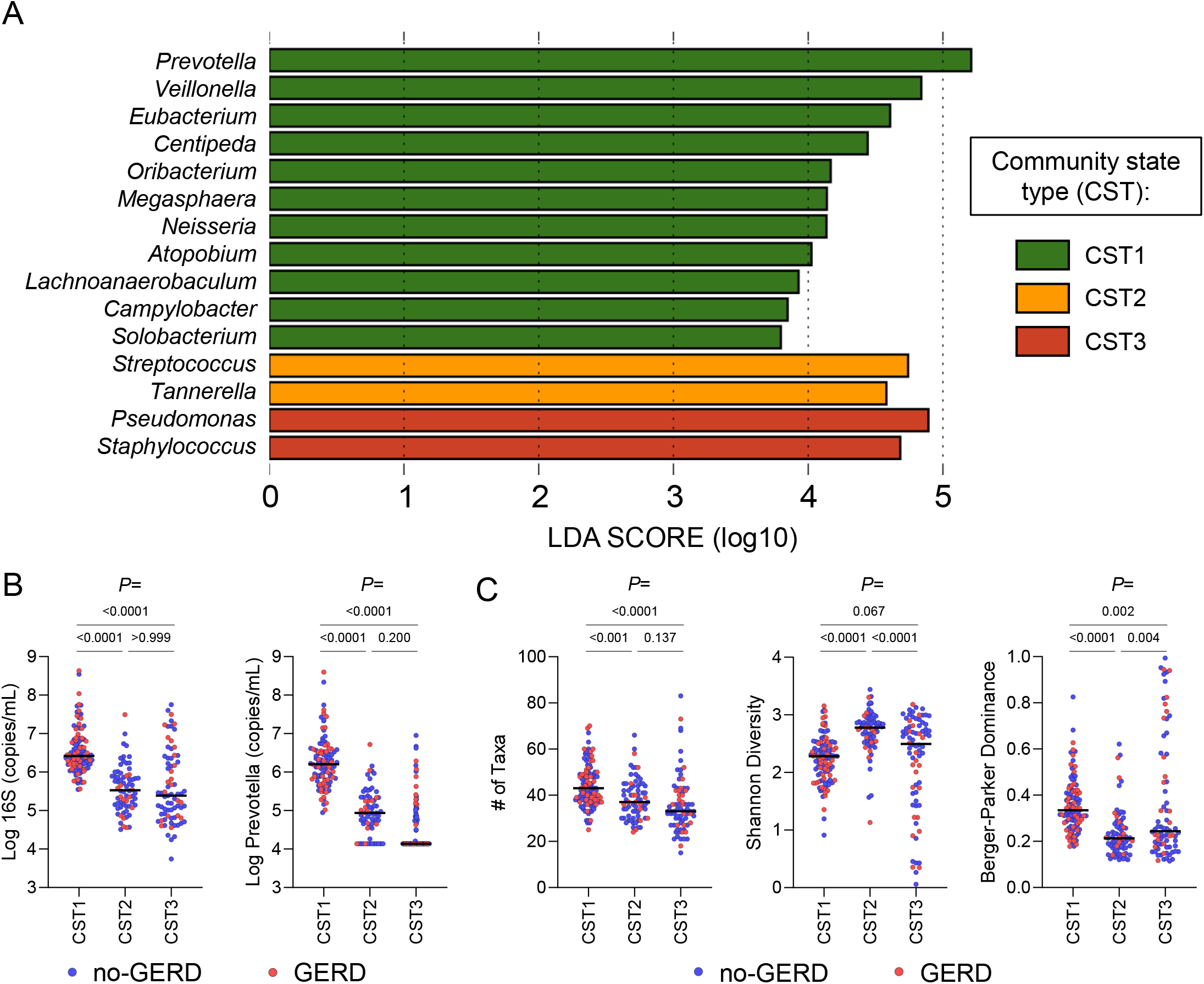
CST-specific microbial features. **A**. Genera enriched in each CST, identified using the LefSe (Linear discriminant analysis (LDA) Effect Size) pipeline. A higher LDA score indicates a higher difference between groups. **B**. Comparison of total bacterial density based on 16S rRNA gene copies/mL BALF and *Prevotella*-specific quantitative polymerase chain reaction, by CST. **C**. Comparison of alpha diversity indices, by CST. Group comparison was tested using Mann-Whitney tests. CST = community state type; BALF = bronchoalveolar lavage fluid.

The median bacterial density (16S rRNA gene copies/mL BALF) was ∼10-fold higher in CST1 than in CST 2 or 3, as was the absolute abundance of the CST1-associated genus *Prevotella* as measured by 16S rRNA gene and *Prevotella*-specific qPCR, respectively (**Figure 2B**). Although taxonomic richness was highest in CST1, composite (Shannon) diversity was lower than CSTs 2 and 3 due to the high relative abundance of *Prevotella*. CST2 had the greatest evenness/lowest tendency towards dominated communities, while CST3 was characterized by high variability in density, diversity and taxonomic dominance, with many samples highly dominated by a single taxon (**Figure 2C**). The most abundant genera in the highly dominated communities in CST3 were *Staphylococcus* and *Pseudomonas*. Thus, CST1 seems to represent a high-bacterial-density state dominated by oropharyngeal taxa, CST2 a low-density state and CST3 a variable-density state commonly characterized by dominance with pathogenic taxa. Although the proportion of samples in each CST differed by GERD status, compositional differences between GERD and no-GERD samples within CSTs were not observed.

#### Longitudinal comparisons in the first post-transplant year

We next assessed whether bacterial density, alpha diversity, CST membership and their longitudinal stability differed between GERD cases and no-GERD controls. Using a generalized estimating equation (GEE) model, we assessed stability in bacterial density over time and observed that GERD patients had significantly greater variability in microbial density than controls (coefficient of correlation for cases: ρ = 0.165, *P* = 0.332 and for controls ρ = 0.153, *P* = 0.01, a *P* < 0.05 indicating stability over time, **Figure 3A**). Shannon diversity and Berger-Parker dominance were consistent over the first year in controls (mean_SDI(no-GERD)_ = 2.35, CI_95_ [2.26-2.45]; mean_BP(no-GERD)_ = 0.33, CI_95_[0.31-0.36]). In GERD patients, both indices were significantly lower than controls at 3 months (MW; mean_SDI(GERD)_ = 1.98, mean_BP(GERD)_ = 0.42, *P* = 0.007 and 0.025, respectively) but not at later timepoints (mean_SDI(GERD)_ (6-12mts) = 2.32, CI95 [2.18-2.48]; mean_BP(GERD)_ (6-12mts) = 0.33, CI95 [0.29-0.36], **Figure 3B**), indicating that recovery of microbial diversity is delayed in patients with GERD.

**Figure 3.**
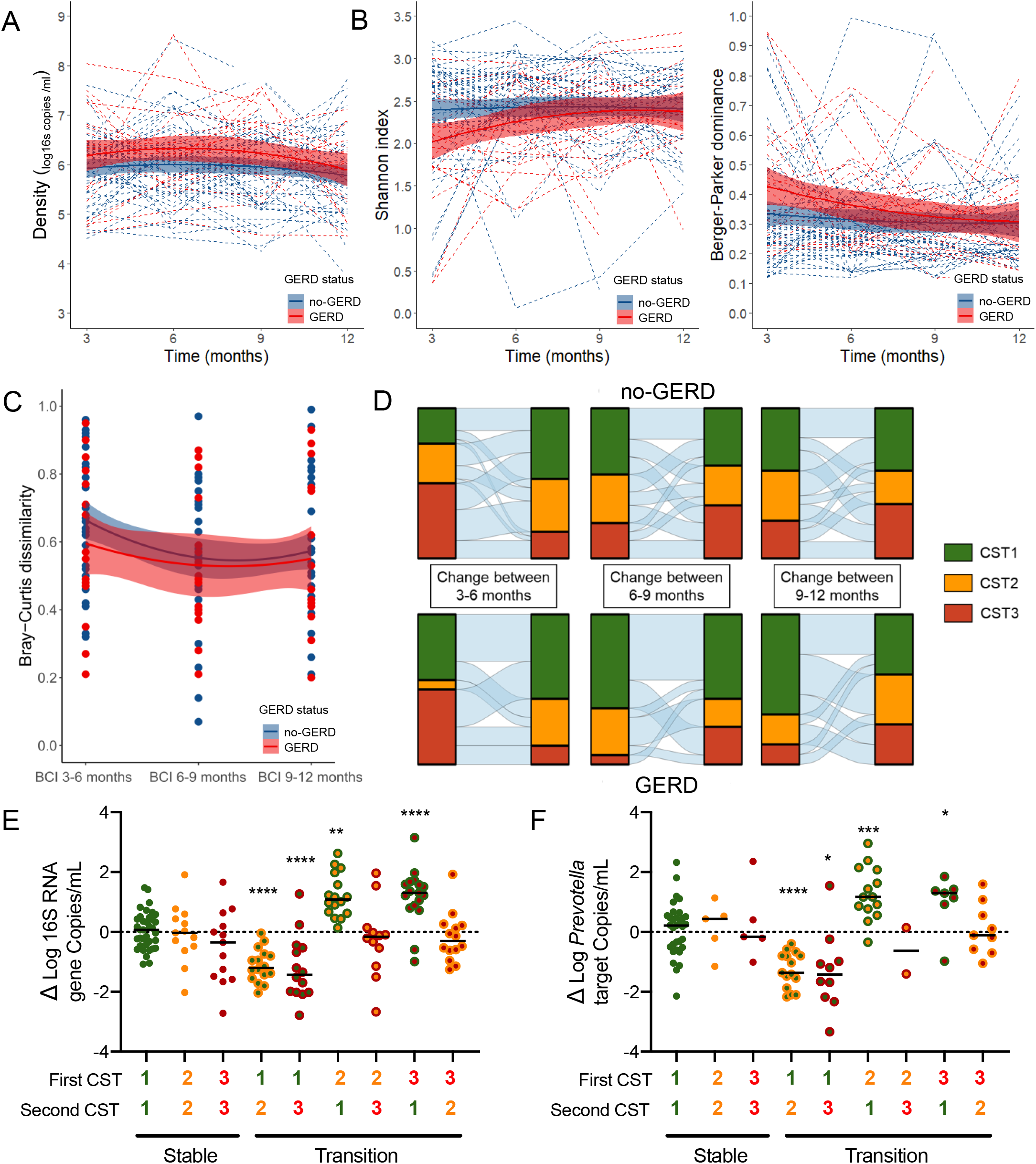
Longitudinal analysis of the lung microbiota 1 year after transplantation. **A**. Comparison of total bacterial density variation by GERD status. **B**. Comparison of alpha diversity indices changes over time by GERD status. **C**. Patient-specific variation measured using the Bray-Curtis dissimilarity measure between consecutive samples, by GERD status. **D**. Transitions of CSTs between 3-6-, 6-9-, and 9-12-months post-transplant, by GERD status. **E**. Changes in total bacterial density associated with transition to the different CSTs. **F**. Changes in *Prevotella* density associated with CST transitions.

Compositional variability over time was greatest in both groups at early sampling intervals (3-6 months) and stabilized over time (**Figure 3C**). While transitions between CSTs were common in both groups, transitions to CST1 were most common in GERD, and CST1 was more stable in GERD patients than controls (**Figure 3D**). Transitions to CST1 were associated with significant increases in absolute total bacterial abundance (**Figure 3E**) and absolute abundance of the CST1-associated genus *Prevotella* (**Figure 3F**). Conversely, transitions from CST1 were associated with significant decreases in absolute total bacterial and *Prevotella* abundance. Transitions between CST 2 and 3 were not associated with changes in absolute bacterial abundance or absolute abundance of *Prevotella*. Instability in bacterial abundance in GERD patients during the first year is thus driven by transitions to and from CST1.

### GERD and CST as predictors of inflammation, ALAD, CLAD, and death

We have previously reported differences in inflammatory cytokine levels, lung allograft dysfunction and GERD in this cohort (1). We sought to assess whether the addition of microbial community composition and density (CST and 16s rRNA gene copy density) affected associations between allograft inflammation, ALAD and CLAD.

#### Inflammation

16S density was strongly associated with individual proinflammatory cytokine levels independently of GERD status (**Figure 4A**). Inflammation was defined as having at least two out of four pro-inflammatory cytokines (IL-1α, IL-1β, IL-6, and IL-8) in the 75^th^ percentile, based on a similar previously published approach (12). Samples with and without inflammation showed distinct distributions of proinflammatory cytokine concentrations (**Figure S4A**). Inflammation was associated with higher bacterial density at 3-, 6-, and 12-months post-transplant (**Figure S4B**). Despite bacterial density being significantly higher in patients with CST1, the proportion of patients with inflammation was significantly higher in patients with CST3 when compared to CST1 (Fisher’s Exact OR [95% CI] = 2.3 [1.2-4.7], *P* = 0.022). Although not significant, this CST3-inflammation relationship held when compared to CST2 (OR [95% CI] = 2.1 [1.0-4.4], *P* = 0.090) as well. There were no differences in the proportion of samples with inflammation from patients with and without GERD within CST1 (Fisher’s Exact OR [95% CI] = 1.6 [0.6-4.0], *P* = 0.462), CST2 (Fisher’s Exact OR [95% CI] = 1.6 [0.5-5.3], *P* = 0.510), and CST3 (Fisher’s Exact OR [95% CI] = 1.4 [0.5-3.9], *P* = 0.596) (**Figure 4B**). Comparison of individual proinflammatory cytokines showed no differences between CSTs, with the exception of CST2 which was associated with lower IL-8 concentrations compared to CST3 (**Figure S4C**). Notably, while bacterial density/inflammation correlations were independent of GERD diagnosis, they differed significantly by CST (**Figure 5**). All four proinflammatory cytokine levels (IL-1α, IL-1β, IL-6 and IL-8) were associated with consistently lower bacterial density for CST3 compared to CST1 (*P*-values for differences of regression intercepts between CSTs were 0.044, 0.014, 0.022 and <0.0001, respectively). This may in part explain why, although GERD is associated with higher and more variable bacterial density in the first transplant year, it is not associated with increased inflammation in our cohort.

**Figure 4.**
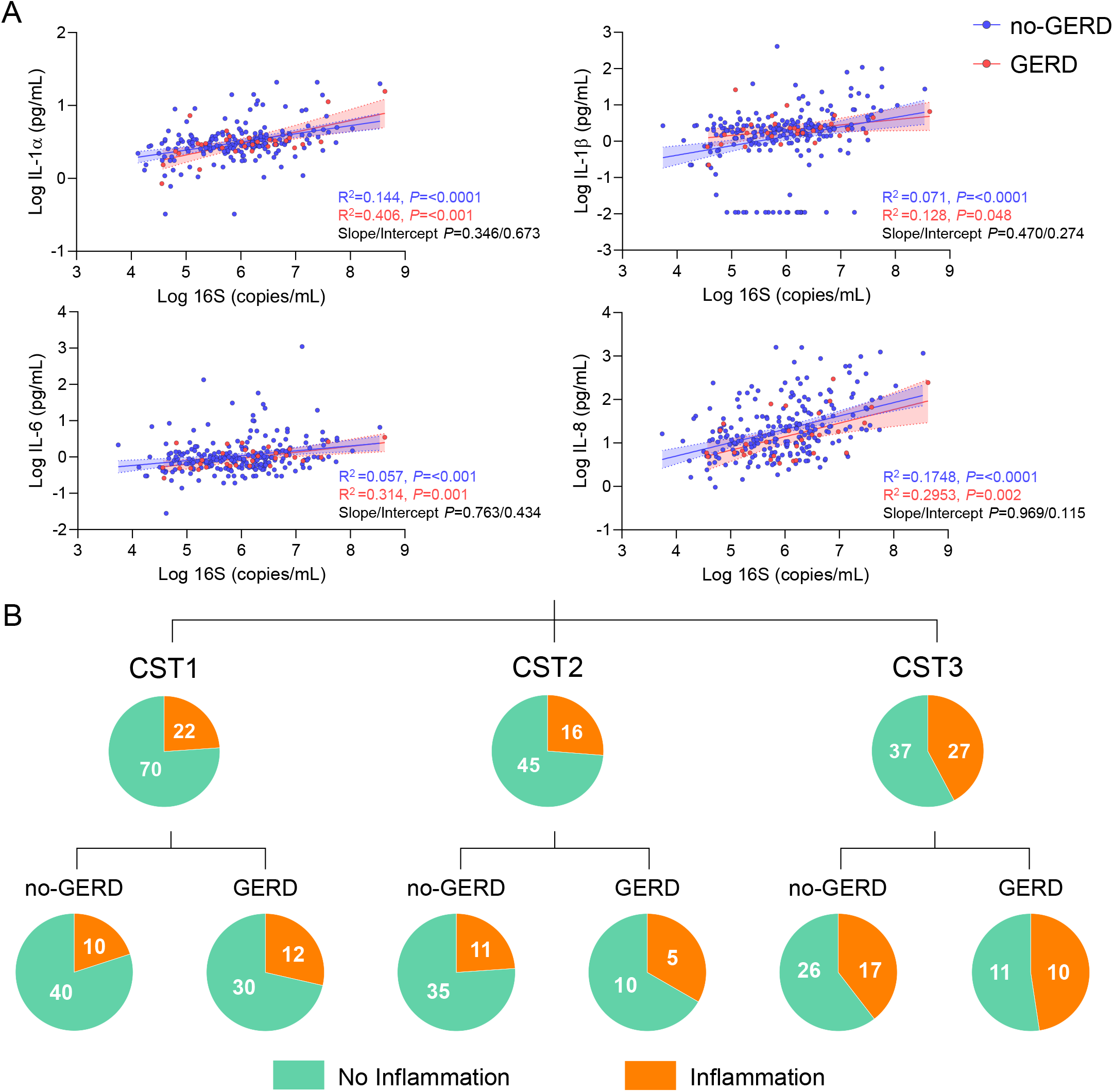
Associations between GERD status, microbial composition and density, and inflammation. **A**. Linear regression analysis of total bacterial density and individual pro-inflammatory cytokines. **B**. Proportion of samples with inflammation - defined by two or more cytokines in the top 75^th^ percentile -, by CST and further stratified by GERD status.

**Figure 5.**
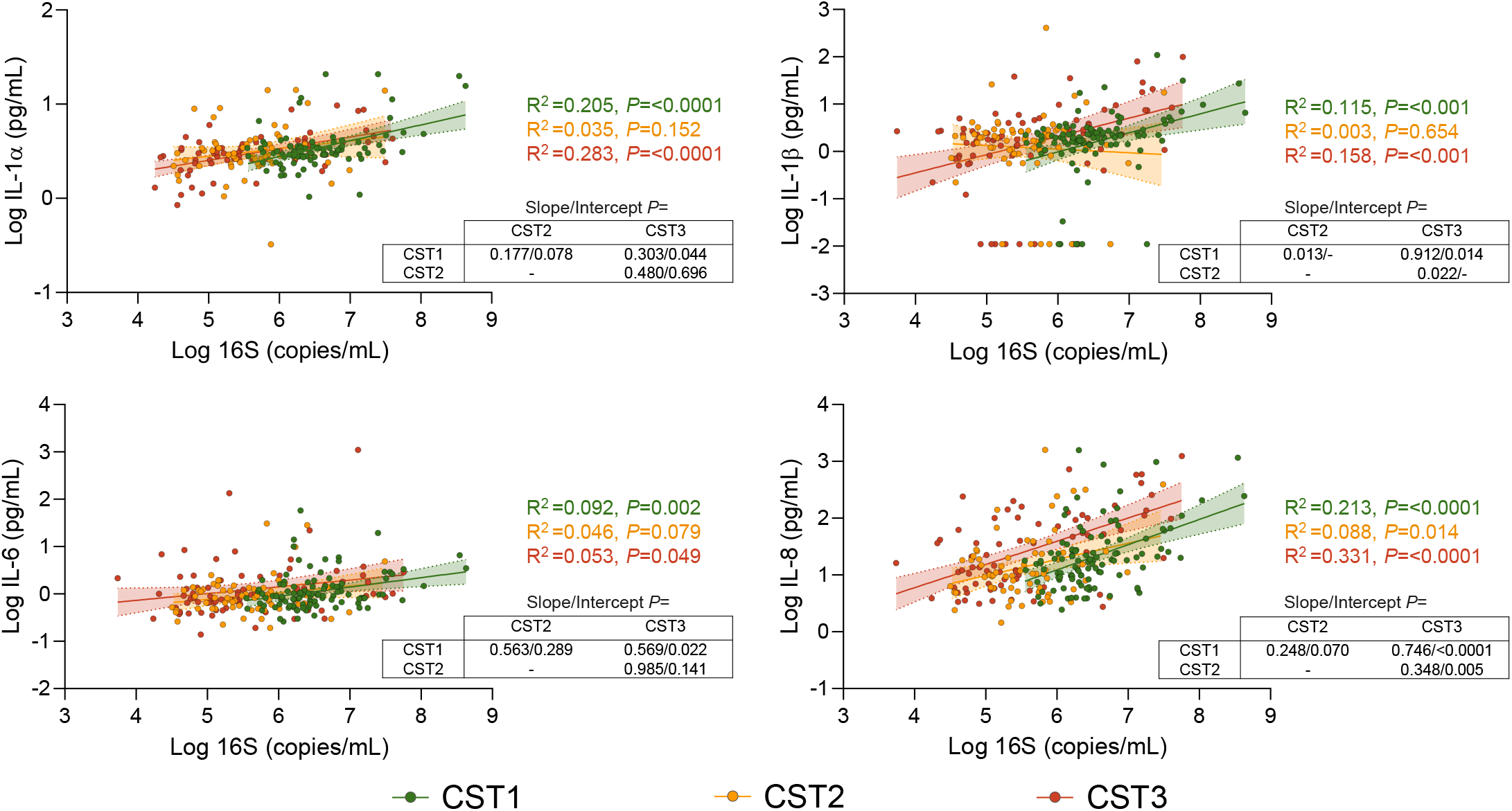
Association between total bacterial density and individual pro-inflammatory cytokines, stratified by community state type (CST1 = green; CST2 = yellow; CST3 = red).

#### ALAD, CLAD and Death

At three months, a diagnosis of GERD was not significantly associated with concurrent ALAD (Fisher’s Exact OR [95% CI] = 1.4 [0.4-6.5], *P* = 0.692). However, ALAD was more common in patients with CST3 than with CST1 (Fisher’s Exact OR [95% CI] = Infinity [1.5-Infinity], *P* = 0.030), but not different from CST2 (Fisher’s Exact OR [95% CI] = 1.8 [0.3-9.3], *P* = 0.694). A Cox proportional hazards model was used to assess the association of GERD status and the number of CST3 events at different intervals with time to CLAD and death, as shown in **Table 3**. One patient was excluded from this analysis due to missing metadata. GERD status was not associated with CLAD. However, patients with higher cumulative number of CST3 events at 6-, 9-, and 12-months post-transplant were more likely to develop CLAD compared to CST1 or CST2, even when adjusted for sex, age at transplant, CMV serostatus mismatch, and primary disease. This relationship also held at 3 months post-transplant when adjusted for sex and CMV mismatch. GERD status and the number of CST3 events were not significantly associated with death.

**Table 3.** Cox proportional hazards associated with GERD and events of community state type 3 (CST3). GERD status and the number of CST3 events at different intervals were used to predict CLAD and death, and adjusted for potential confounders. HR = hazard ratios; G+ = GERD positive; mts = months; EV = events; tx = transplant; CMV = cytomegalovirus

| Outcome | Predictor | HR (95% CI) |  | HR (95% CI) |  | HR (95% CI) |  | HR (95% CI) |  | HR (95% CI) |  |
| --- | --- | --- | --- | --- | --- | --- | --- | --- | --- | --- | --- |
|  |  | GERD status (GERD) | P | CST3 samples at 3 mts | P | CST3 samples up to 6 mts | P | CST3 samples up to 9 mts | P | CST3 samples up to 12 mts | P |
| CLAD |  | n=74, EV=21 |  | n=67, EV=20 |  | n=72, EV=19 |  | n=70, EV=19 |  | n=69, EV=18 |  |
|  |  | 0.55<br>(0.20-1.51) | 0.24 | 1.99<br>(0.82-4.87) | 0.12 | 2.01<br>(1.08-3.75) | 0.02 | 1.77<br>(1.05-2.97) | 0.03 | 1.67<br>(1.01-2.74) | 0.04 |
|  | Adjusted for |  |  |  |  |  |  |  |  |  |  |
|  | Sex (M) | 0.53<br>(0.19-1.46) | 0.22 | 2.43<br>(0.96-6.13) | 0.06 | 2.56<br>(1.29-5.07) | 0.01 | 2.11<br>(1.21-3.68) | 0.01 | 1.76<br>(1.08-2.85) | 0.02 |
|  | Age at tx | 0.41<br>(0.14-1.20) | 0.11 | 1.87<br>(0.76-4.61) | 0.17 | 1.88<br>(0.98-3.63) | 0.05 | 1.68<br>(0.97-2.91) | 0.06 | 1.57<br>(0.95-2.61) | 0.07 |
|  | CMV mismatch (Yes) | 0.55<br>(0.20-1.51) | 0.24 | 2.20<br>(0.87-5.53) | 0.09 | 2.05<br>(1.10-3.82) | 0.02 | 1.76<br>(1.05-2.97) | 0.03 | 1.67<br>(1.01-2.78) | 0.04 |
|  | Primary disease | 0.46<br>(0.16-1.29) | 0.14 | 1.89<br>(0.75-4.76) | 0.17 | 1.82<br>(0.96-3.46) | 0.06 | 1.56<br>(0.93-2.62) | 0.09 | 1.55<br>(0.94-2.54) | 0.08 |
| DEATH |  | n=74, EV=19 |  | n=67, EV=17 |  | n=72, EV=17 |  | n=70, EV=15 |  | n=69, EV=14 |  |
|  |  | 0.37<br>(.11-1.28) | 0.12 | 1.30<br>(0.50-3.38) | 0.59 | 1.35<br>(0.66-2.73) | 0.41 | 1.21<br>(0.66-2.22) | 0.54 | 1.26<br>(0.72-2.23) | 0.42 |
|  | Adjusted for |  |  |  |  |  |  |  |  |  |  |
|  | Sex (M) | 0.36<br>(0.11-1.26) | 0.11 | 1.51<br>(0.57-4.04) | 0.41 | 1.50<br>(0.71-3.19) | 0.29 | 1.31<br>(0.70-2.44) | 0.39 | 1.29<br>(0.75-2.26) | 0.36 |
|  | Age at tx | 0.36<br>(0.10-1.29) | 0.12 | 1.29<br>(0.49-3.40) | 0.6 | 1.49<br>(0.69-3.22) | 0.31 | 1.25<br>(0.66-2.35) | 0.49 | 1.27<br>(0.71-2.26) | 0.41 |
|  | CMV mismatch (Yes) | 0.37<br>(0.11-1.29) | 0.12 | 1.49<br>(0.55-4.04) | 0.43 | 1.43<br>(0.72-2.86) | 0.31 | 1.23<br>(0.68-2.21) | 0.49 | 1.28<br>(0.74-2.21) | 0.37 |
|  | Primary disease | 0.36<br>(0.10-1.24) | 0.11 | 1.22<br>(0.46-3.25) | 0.7 | 1.51<br>(0.72-3.16) | 0.27 | 1.24<br>(0.68-2.27) | 0.48 | 1.24<br>(0.72-2.11) | 0.44 |

### Association of Nissen Fundoplication with Inflammatory cytokines and bacterial density

In a cohort of 10-patients who underwent Nissen fundoplication for symptomatic GERD, we assessed pre/post-procedure bacterial load and pro-inflammatory cytokine levels in BALF supernatants. Similarly to our previous report (1), significant decreases in pro-inflammatory cytokines were observed after fundoplication (**Figure 6A**). While no significant changes were observed in overall bacterial loads pre and post fundoplication, individuals with the greatest decreases in inflammatory cytokines also had the greatest decreases in bacterial density as measured by 16S rRNA gene qPCR, and had the highest baseline densities of the CST1-associated genus *Prevotella*. This suggests that fundoplication may have direct or indirect effects on the allograft microbiota that vary by baseline community composition (**Figure 6B**).

**Figure 6.**
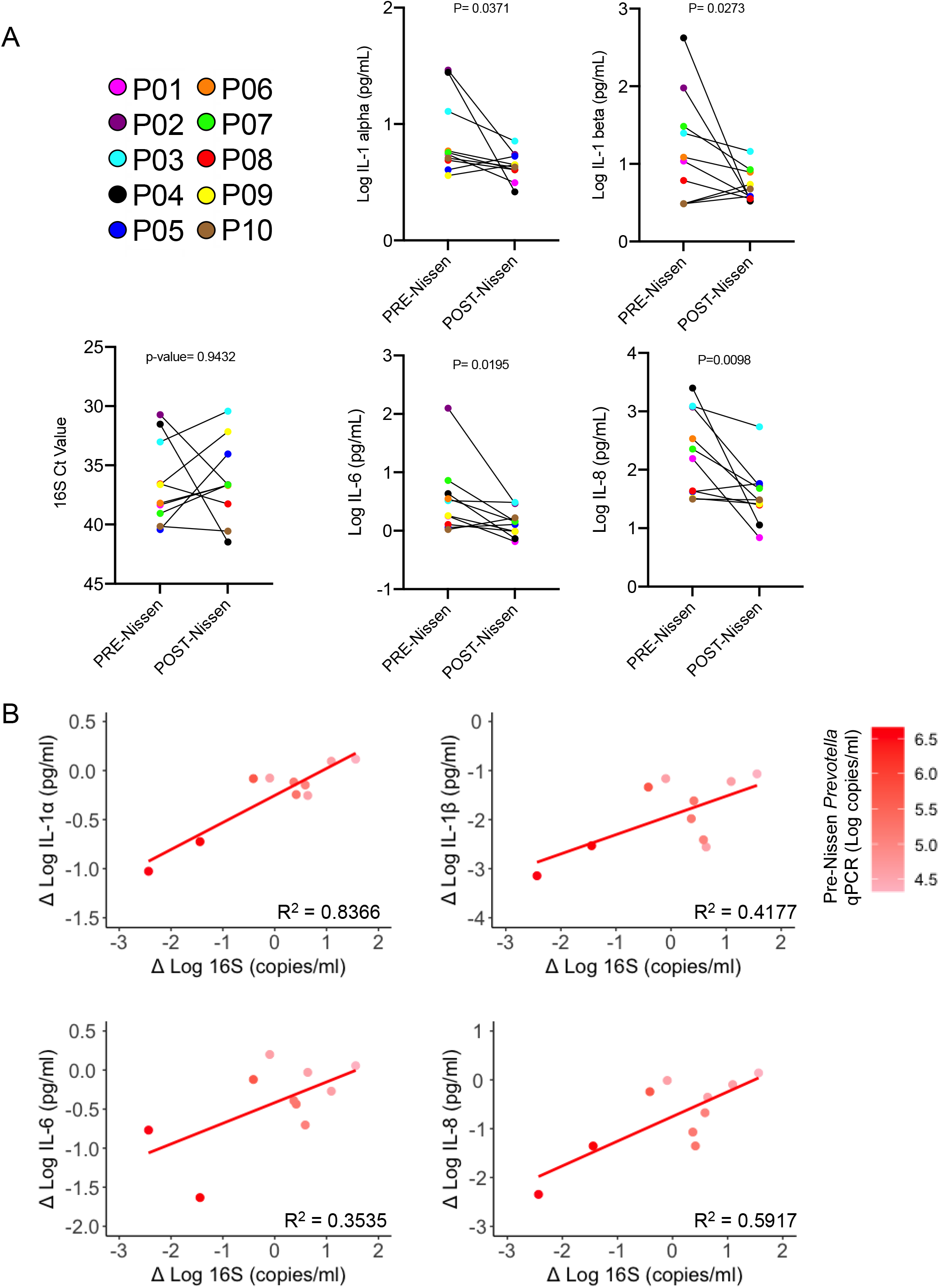
Pre/Post-Nissen fundoplication comparison of bacterial density and individual cytokines. **A**. Changes in total bacterial density and cytokines, by patient. **B**. Regression between individual cytokines and total bacterial density, weighted by pre-Nissen density of the CST1-associated genus, *Prevotella*.

## Discussion

This study is the first to systematically compare the allograft microbiota in lung transplant recipients with and without GERD. We were able to compare the relationship between microbial community composition, inflammation, ALAD and CLAD in lung allograft recipients with and without GERD, while also assessing the effects of anti-reflux surgery on allograft bacterial density. We believe we add the following observations to our understanding of allograft microbiota-host associations in the context of GERD:

1. The strongest feature of allograft community composition is the underlying compositional clustering of the allograft microbiota. We observed three major clusters, which we referred to as CST 1-3, with no significant observed microbial community compositional differences between GERD cases and controls within these clusters;
2. The proportion of samples with these CSTs differed between GERD and control groups, with the high-density, oropharyngeal-taxa-enriched CST1 being more common in GERD than in no-GERD controls;
3. GERD was associated with a more variable community composition and bacterial abundance in the first-year post-transplant, with more frequent transitions between lower density CST2/3 and CST1 in GERD cases than controls;
4. CST-inflammation associations exceeded any association between GERD status and inflammation in this cohort;
5. The CST most common in lung allograft recipients with GERD (CST1) was associated with lower per-bacteria inflammatory cytokine levels than the pathogen-dominated CST3;
6. Models used to predict inflammation, ALAD and CLAD revealed CST – particularly CST3, but not GERD-status associations;
7. Nissen fundoplication was associated with strongly correlated decreases in bacterial load and pro-inflammatory cytokines, especially in individuals with high pre-fundoplication density of the CST1-associated genus *Prevotella*.

Our observation of a clear structure underlying beta-diversity of the lung allograft confirms the results of others (5, 8). We applied the term ‘community state types’ to describe these distinct clusters as is commonly done to describe the microbiota of the female genital tract (13), which has a similarly limited number of taxa and discrete compositional clustering compared to other human sample types such as gut/stool. We note that there are taxonomic differences in the clusters we observed compared to those reported by others, which may in part be due to technical factors (e.g. sequencing methods and sequence variant annotation) or differences in methods to identify and remove taxa suspected to be reagent contaminants, which is especially consequential for low density samples in CST2 (14). Nonetheless, we noted a very consistent composition for CST1, dominated by oropharyngeal taxa and CST3 characterized by the prevalence and relative abundance of putative pathogens. Thus, we feel our findings with respect to community composition are likely to be generalizable to other cohorts.

We were unable to assess whether enrichment for CST1 amongst individuals with GERD was due to actual aspiration of refluxed oropharyngeal content in this observational study. However, we found that decreases in inflammatory cytokines after Nissen fundoplication were greatest in individuals with significant decreases in bacterial density and high initial *Prevotella* absolute abundance in BALF supernatant. This is consistent with a model in which GERD is associated with aspiration of oropharyngeal taxa, leading to inflammation. This hypothesis requires both replication and experimental validation.

CST was not significantly associated with bile acid concentrations, although we cannot exclude the possibility that this association exists and future datasets can reassess this in larger cohorts. Conjugated bile acids, like TCA and GCA, have been associated with poor clinical outcomes and bacterial infections (1, 15). This may explain the weak trends in CST1 and CST3, CSTs that contain aspiration-related microbes and pathogens.

A similar number of patients with and without GERD had CST3, suggesting that this CST arises independently of reflux. CST3 was characterized by increased relative abundance of *Pseudomonas* and *Staphylococcus*, two taxa with known pathogenic species. Despite CST1 being enriched for GERD and having higher bacterial density, CST1 and CST2 were similarly associated with decreased rates of inflammation, CLAD, and ALAD compared to CST3. The association of pathogen-communities, inflammation and host status replicates the findings of others (5, 8). Importantly, our study highlights how the relationship between GERD and clinical outcomes may be confounded by the underlying microbial composition of the lung, since the GERD-associated CST1 correlated with less inflammation, ALAD and CLAD. Future studies assessing relationships between GERD and host status or prognosis should control for the potential confounding by underlying CST or analyse it as an interacting variable.

Our study has several important limitations. The primary analysis was based on a case-control, retrospective, single-centre cohort, requiring validation. In addition, the cohort was composed of patients with clear GERD or no-GERD testing results, including individuals with numbers of reflux episodes in 24 hours of >48 or <23. Our findings, therefore, may not apply to individuals with intermittent or less frequent/severe reflux. Importantly, this limitation would be expected to exaggerate the differences between cases and controls, and thus our observation that underlying CST structure is more strongly associated with host status and CLAD than GERD is likely true, as our cohort represents an ‘extreme’ GERD phenotype. In addition, we did not comprehensively assess the relationships between CST, GERD and a wide array of soluble or other host factors in BALF, and restricted our analysis to four pro-inflammatory cytokines, ALAD and CLAD. Associations between GERD and other soluble or host-derived factors in BALF may be present. Finally, our small Nissen fundoplication cohort was assessed without sequencing, since we had only BALF supernatant available, which is compositionally different from BALF pellets or raw BALF (uncentrifuged or fractionated).

In summary, increased bacterial density and taxonomic composition dominated by oropharyngeal taxa is associated with GERD. This microbial state was not associated with increased risk of CLAD or death and surgical treatment of GERD showed correlation between changes in bacterial density and levels of pro-inflammatory cytokines. Similarly to what was recently published, we showed that a pathogen-dominated community independent of GERD status is associated with increased risk of lung allograft dysfunction. Future studies should investigate the exact effect of GERD treatment and whether it promotes or protects from colonization of the lung ecological niche by CLAD-associated microbial communities.

## Methods

### Cohort design

This study was approved by the Toronto University Health Network Research Ethics Board (REB#15-9698-AE) and the Duke University Internal Review Board (Pro00013378). The cohorts were derived from two previously published cohorts, leveraging the cytokine and bile acid data that was generated by this prior study (1).

#### GERD Cohort (Figure S1)

Patients from the previously published GERD cohort were included if they had at least one available raw (unprocessed) BALF sample obtained in the first-year post-transplant (1). In summary, this was a nested case-control cohort, drawn from all adults who underwent bilateral or single lung transplantation at Toronto General Hospital between 2010 and 2015. Patients were included if they had 24-hour pH/impedance probe testing and at least one raw and supernatant BALF sample collected and preserved in the first-year post-transplant. Patients whose 24-hour pH/impedance study showed ≥48 reflux episodes were included as GERD cases. For no-GERD controls, patients with < 23 reflux episodes were excluded if they reported symptoms of reflux during the study or had underlying disease commonly associated with GERD (e.g. scleroderma, hiatal hernia, Barrett’s esophagus, history of Nissen fundoplication or pyloroplasty), and matched 2:1 to the GERD patients by transplant type (single vs. bilateral). For the selected patients, available BALF samples closest to the 3-, 6-, 9-, and 12-month time points of surveillance bronchoscopies were obtained.

#### Nissen Cohort

Patients from the previously published Nissen cohort were included if they had sufficient pre- and post-Nissen BALF supernatant remaining for analysis (1). In summary, this was a retrospective cohort drawn from all adults who underwent bilateral lung transplantation at Duke University Medical Center between 2005 and 2008. 10 patients were included who underwent Nissen fundoplication within 6 months post-transplant and had available paired BALF samples obtained within 3 months before and after Nissen fundoplication.

### Clinical Standards of Care and Definitions

Standard of care for lung transplant recipients was delivered as described previously by the Toronto and Duke programs (16, 17). In the first year after lung transplant, both programs performed surveillance bronchoscopies to obtain BALF and transbronchial biopsies at pre-specified time points (Toronto: 0.5, 1.5, 3, 6, 9, 12 months; Duke: 1, 3, 6, 9, 12 months) and as clinically indicated. During the study period, the Toronto BAL protocol was to instill 50 mL of normal saline twice, while the Duke protocol consisted of one 60 mL instillation, the right middle lobe being the default location at both institutions.

At Toronto, patients were treated with proton pump inhibitors based on symptoms and routine pH/impedance studies performed at 3 months post-transplant. No patients in the cohort underwent Nissen fundoplication in the first post-transplant year.

At Duke, patients were treated with proton pump inhibitors regardless of reflux studies. Patients underwent routine 24-hour pH monitor prior to transplantation, and repeat testing post-transplant if the pre-transplant study was negative. Any positive study, defined as <0.9% proximal acid contact time and/or <4.2% distal acid contact time, led to referral for Nissen fundoplication.

Acute lung allograft dysfunction (ALAD) was defined as a ≥10% decline in measured forced expiratory volume in 1 second (FEV1) compared to the higher of the two preceding FEV1 measurements. CLAD was defined as per latest consensus report from the International Society for Heart and Lung Transplantation (2).

### BAL sample processing

In Toronto, raw BALF samples were aliquoted and stored at -80 °C. The remaining BALF was centrifuged at 3184 g for 20 minutes and the supernatant was also stored at -80 °C. At Duke, BALF samples were centrifuged at 1750 g for 10 minutes at 4°C and the supernatant was stored at -80 °C.

### Analysis of BALF supernatant samples

Markers of innate immune activation, including IL-1α, IL-1β, IL-6, and IL-8, were measured in the BALF supernatant using a custom-designed multiplex assay (R&D), as reported previously (1). Taurocholic acid (TCA), one of the most abundant bile acids, was measured using LC-MS/MS mass spectrometry, as reported previously (1).

### DNA isolation and quantification from BALF samples

Nucleic acids were isolated from 250 µl of raw BALF samples (Toronto) or BALF supernatant (Duke) using a PowerSoil DNA isolation kit (MO-BIO; Carlsbad, CA, USA) following the manufacturer’s instructions except for the elution step which was done in 60 µl purified water. Densities were measured using a 16S quantitative polymerase chain reaction (qPCR;(18)) and a standard (*Pseudomonas aeruginosa* str. PAO1), and the number of 16S copies /ml was inferred using the URI Genomics & Sequencing Center online calculator (http://cels.uri.edu/gsc/cndna.html). 16S qPCR primers and conditions are described in the online supplement. qPCR reactions were carried out in a volume of 11 µl using the TaqMan Gene Expression Master Mix (Applied Biosystems, Foster City, CA, USA) according to the manufacturer’s protocol.

### 16S rRNA gene sequencing of DNA from raw BALF samples

The V4 hypervariable region of the 16S rRNA gene is amplified using a universal forward sequencing primer and a uniquely barcoded reverse sequencing primer to allow for multiplexing (19). Amplification reactions are performed using 12.5 µL of KAPA2G Robust HotStart ReadyMix (KAPA Biosystems), 1.5 µL of 10 µM forward and reverse primers, 7.5 µL of sterile water and 2 µL of DNA. The V4 region was amplified by cycling the reaction at 95°C for 3 minutes, 28x cycles of 95°C for 15 seconds, 50°C for 15 seconds and 72°C for 15 seconds, followed by a 5-minute 72°C extension. All amplification reactions were done in triplicate, checked on a 1% agarose TBE gel, and then pooled to reduce amplification bias. Pooled triplicates were quantified using PicoGreen (Thermo Fisher) and combined by even concentrations. The library was then purified using Ampure XP beads and loaded on to the Illumina MiSeq for sequencing, according to manufacturer instructions (Illumina, San Diego, CA). Sequencing is performed using the V2 (150bp x 2) chemistry. DNA from a single-species, *Pseudomonas aeruginosa*, from a mock community (Zymo Microbial Standard: https://www.zymoresearch.de/zymobiomics-community-standard), and a template-free negative control were included in the sequencing run. Two individual libraries were prepared and sequenced from each sample.

### Analysis of the bacterial microbiome

The UNOISE pipeline, available through USEARCH v11.0.667 and vsearch v2.10.4, was used for sequence analysis (20-22). The last base was removed from all sequences using cutadapt v.1.18. Sequences were assembled and quality trimmed using –fastq_mergepairs with a –fastq_trunctail set at 5, a –fastq_minqual set at 5, and minimum and maximum assemble lengths set at 243 and 263 (+/-10 from the mean) base pairs. Sequences were first de-replicated and sorted to remove singletons, then denoised and chimeras were removed using the unoise3 command. Assembled sequences were mapped back to the chimera-free denoised sequences at 97% identity OTUs. Taxonomy assignment was executed using SINTAX (23) available through USEARCH, and the UNOISE compatible Ribosomal Database Project (RDP) database version 16, with a minimum confidence cutoff of 0.8 (24). OTU sequences were aligned using align_seqs.py v.1.9.1 through QIIME1 (25). Sequences that did not align were removed from the dataset and a phylogenetic tree of the filtered aligned sequence data was made using FastTree (26).

### Post-profiling filtering approaches

16S rRNA gene sequences from contaminants is a recurrent issue when analysing BALF samples (27) and we controlled for sequencing contaminants as described in Schneeberger *et al*. (14). Briefly, OTUs which were negatively correlated with bacterial density measured with qPCR across all samples were considered as contaminants. In addition, OTUs which were only present in one sequencing replicate were removed.

### Statistical analysis

Bray-Curtis dissimilarity indices were calculated using the “dissimilarity” function from the Vegan R package version 2.5-2 (28). The metaMDS function was used with the options “k=6” and “try=500” to generate NMDS ordination plots. Most important features for CST classification were identified using the random forest model from the randomForest package version 4.6-14 based on (29) and coordinates from the most important classification features (= genus) were extracted manually from the metaMDS output. The correlation coefficients assessing stability in bacterial density were estimated by generalized estimating equations (GEE) from the geepack R package version 1.3-2 (30). Odds ratios and independence were calculated using the Fisher’s exact test. Taxonomic differences between CSTs were identified using the LEfSe pipeline (31). Mann-Whitney tests, Wilcoxon signed rank tests, and Spearman correlations were calculated using XLSTAT 2019 (Addinsoft: Paris, France). Regression analyses were conducted in Graphpad Prism version 9.0.0 (San Diege, CA, USA). Plots were generated using OriginPro 2017 (Northampton, MA, USA) and the R packages ggplot2 version 3.0.0 (32), reshape2 version 1.4.3 (33), and ggalluvial version 0.12.1 based on (34).

## Data Availability

Sequence data that support the findings of this study have been deposited in the NCBI Short Read Archive with the primary accession code PRJNA754787.

## Author contributions

**PHHS:** research design, experiments (microbial culture, pre-sequencing workup, concentration, DNA isolation, 16S qPCR quantification, taxonomic profiling), statistical analyses, figures generation, writing of the initial manuscript, manuscript editing; **CYKZ**: cohort design, cohort organization, clinical data collection, patient phenotyping, sample organization, multiplex data analysis, writing and editing manuscript; **JS**: statistical analyses, figures generation, writing of the initial manuscript, manuscript editing; **BC** and **WX**: statistical analyses, manuscript editing; **YL**: experiments (pre-sequencing workup, DNA isolation, qPCR), manuscript editing; **ZW**: sample organization, experiments for Nissen cohort (DNA isolation, 16S qPCR quantification), qPCR data analysis; **ERN** and **NY**: sample processing, manuscript editing **MA**: cohort design, cohort organization, clinical data collection, patient phenotyping; **KB**: Multiplex cytokine assays; **RR**: clinical data collection and curation; **CWF**: cohort organization, clinical data collection, sample organization (for Nissen cohort); **SMP** and **JLT**: cohort design, clinical data collection, patient phenotyping; **TM**: research design, project supervision, cohort design, data analysis, manuscript writing and editing; **BC**: research design, project supervision, data analysis, manuscript writing and editing.

## Acknowledgements

The authors thank the Centre for the Analysis of Genome Evolution and Function (CAGEF) for providing support in the sequencing experiments as well as bioinformatics support for the taxonomic profiling. The authors also thank the Toronto Lung Transplant Program Biobank team for helping with sample collection and retrieval.

---

**FIGURE S1.**
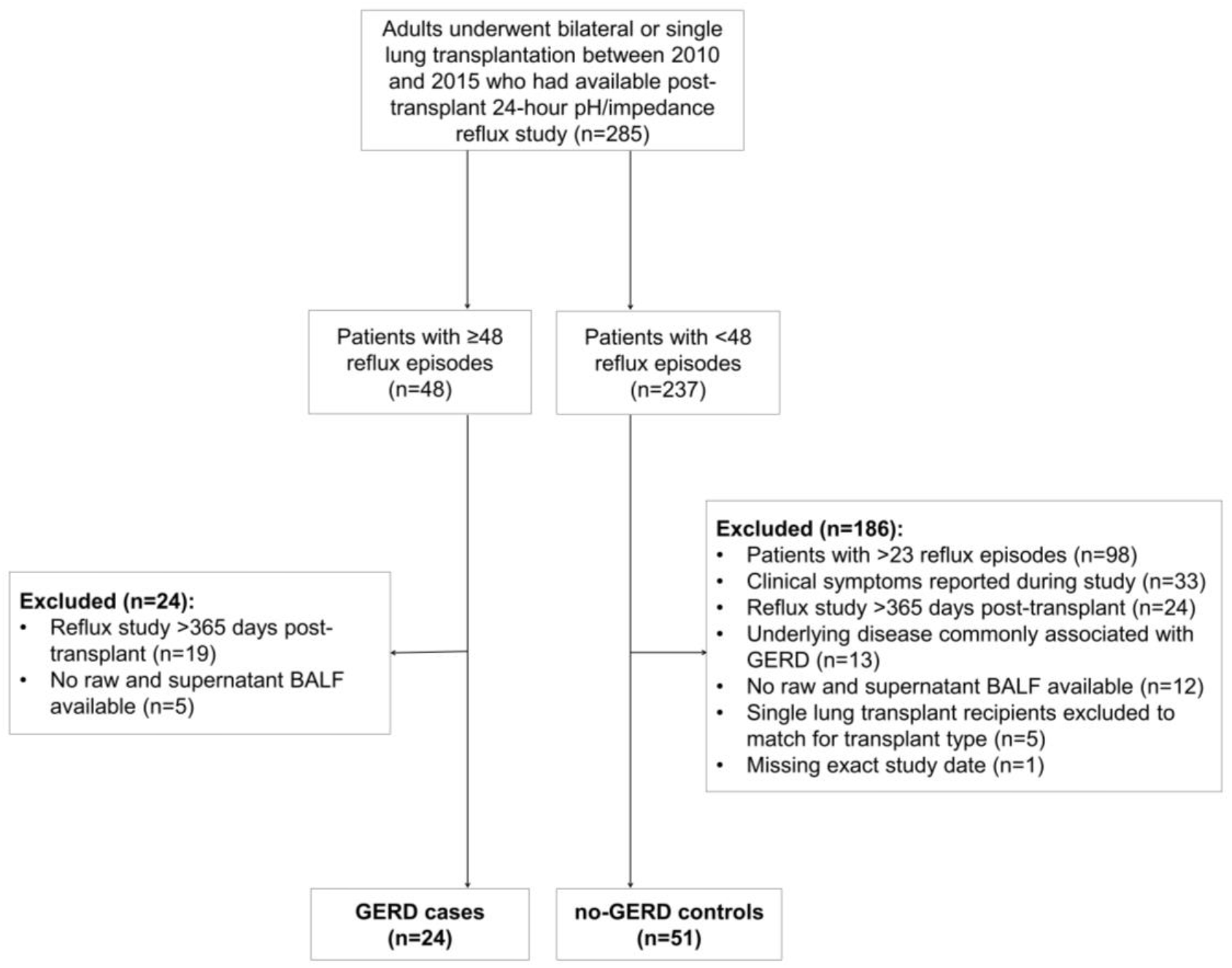

**FIGURE S2.**
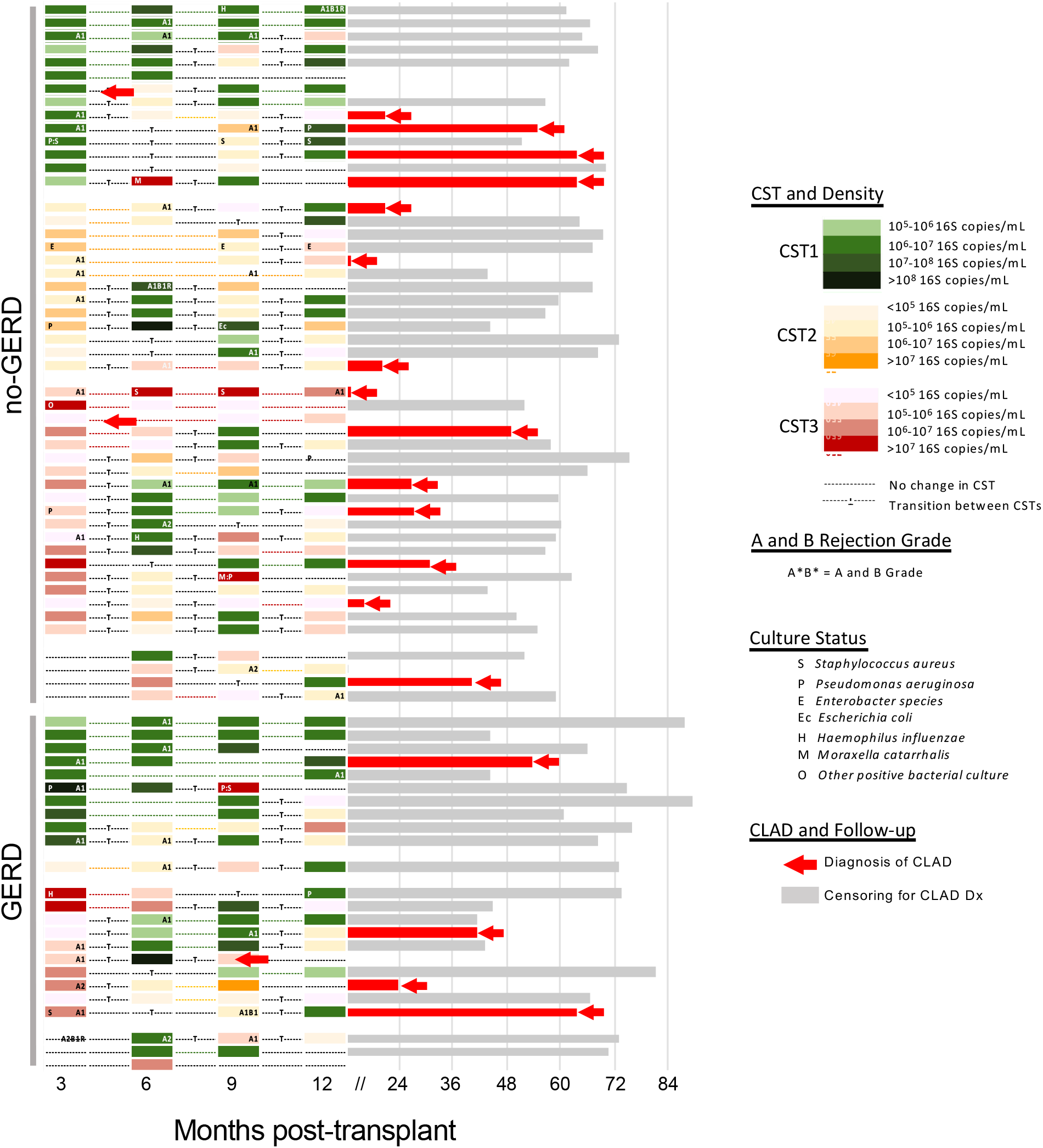

**FIGURE S3.**
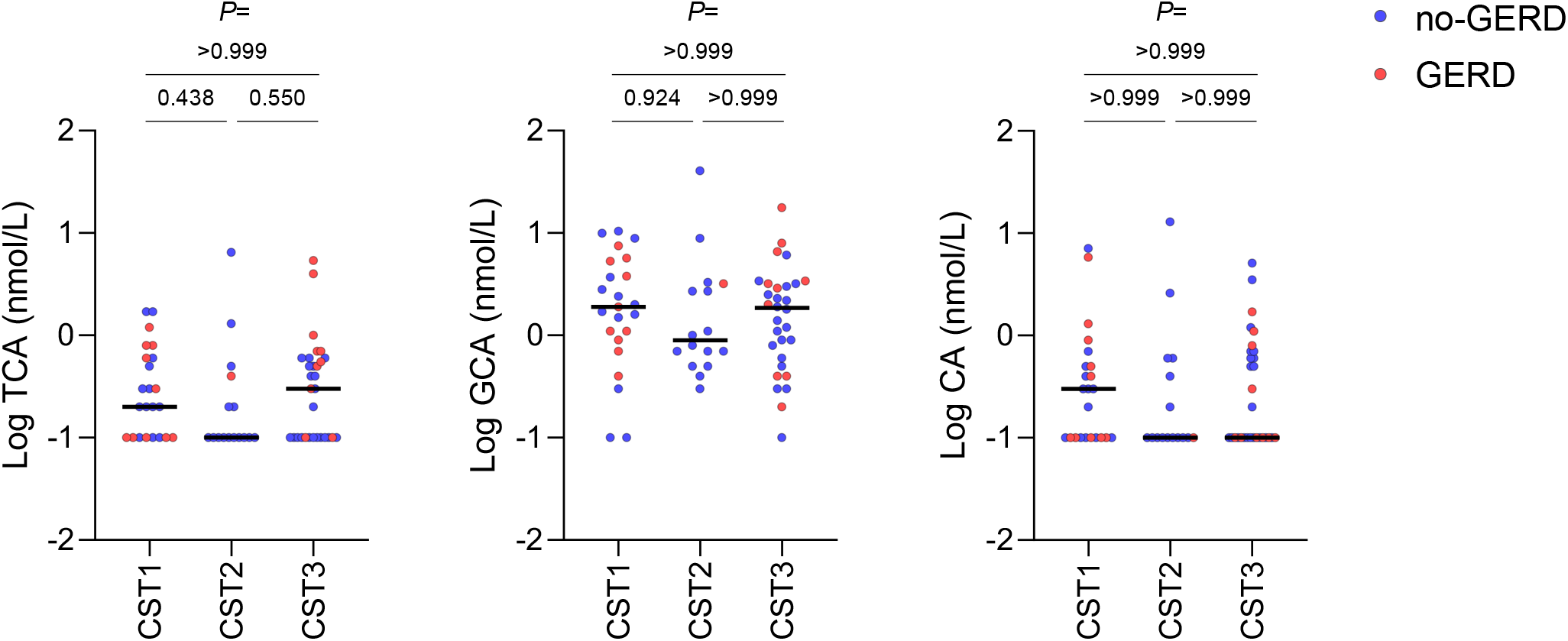

**FIGURE S4.**
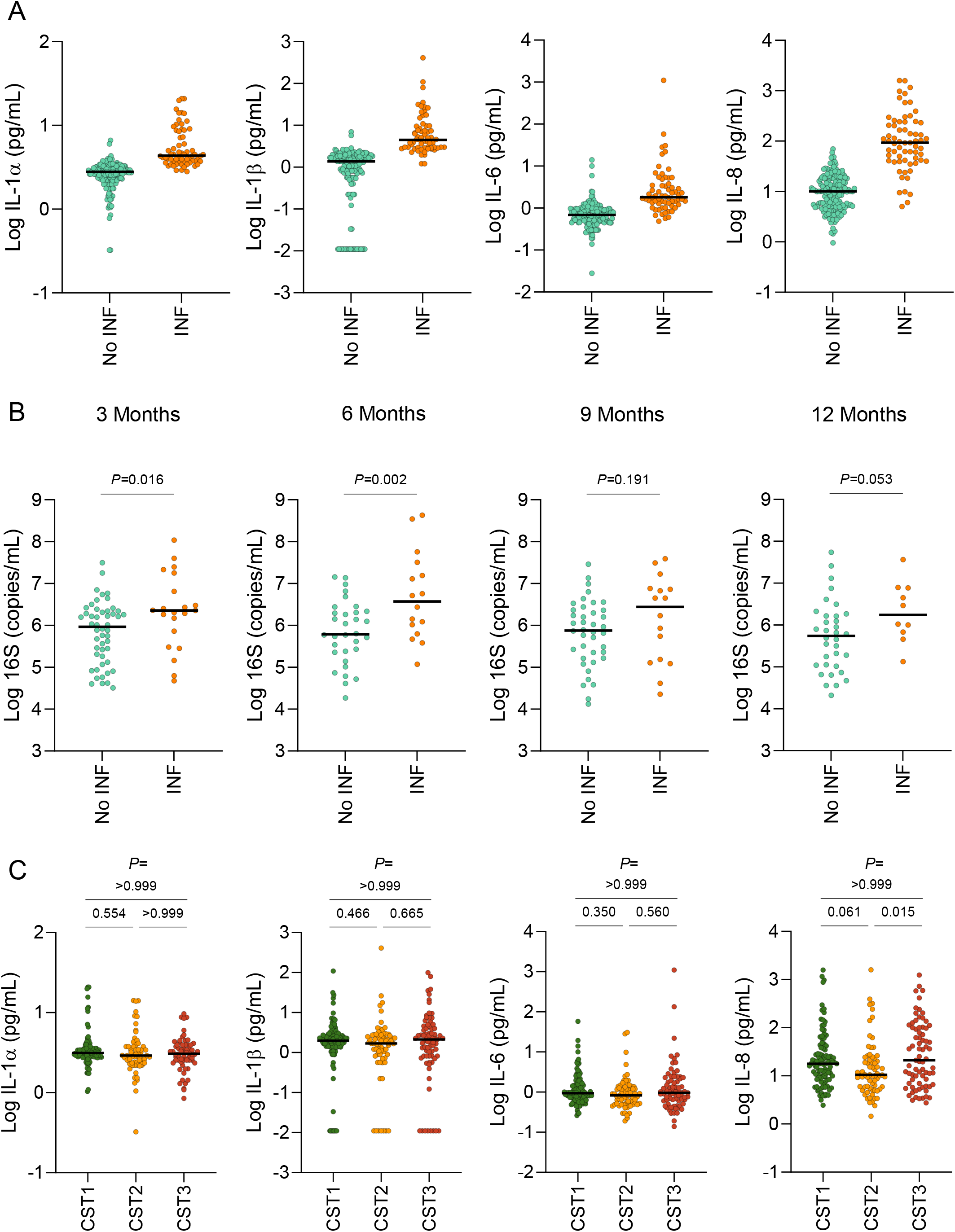

